# Metabolic Dysfunction-associated Steatotic Liver Disease Exacerbates Heart Failure Risk in Patients with Chronic Kidney Disease: A Nationwide Cohort Study

**DOI:** 10.1101/2025.10.21.25338502

**Authors:** Han Na Jang, Kyung-Do Han, Seyun Kim, Sun Joon Moon, Eun-Jung Rhee, Won-Young Lee

## Abstract

**Background:** Steatotic liver disease (SLD) and chronic kidney disease (CKD) are known risk factors for heart failure (HF). We aimed to investigate the risk of developing HF associated with SLD in patients with CKD.

**Methods:** Individuals aged ≥ 20 years who underwent health checkups in 2012 were analyzed using data from the National Health Insurance Services of South Korea. Patients with CKD and an estimated glomerular filtration rate < 60 mL/min/1.73m^2^ or a previous diagnosis of end-stage renal disease were classified into the following categories for comparison of occurrence of HF: no steatosis, metabolic dysfunction-associated steatotic liver disease (MASLD), MASLD with other combined etiology, MASLD and increased alcohol intake, alcohol-associated liver disease, specific etiology SLD, cryptogenic SLD. Liver steatosis was defined as fatty liver index (FLI) ≥ 30.

**Results:** A total of 169,500 patients with CKD were analyzed, 42.4% were male, the mean age was 62.8 ± 12.6 years, and the mean body mass index was 24.4 ± 3.3 kg/m^2^. MASLD (hazard ratio [95% confidence interval]: 1.133 [1.106, 1.161]), MASLD with other combined etiology (1.276 [1.207, 1.350]), and cryptogenic SLD (1.817 [1.095, 3.015]) were associated with higher incidence of HF than those without steatosis. Additionally, an increment of FLI and cardiometabolic risk factors were associated with an increased risk of HF.

**Conclusions:** The incidence of HF was higher in patients with CKD who also had MASLD or cryptogenic SLD. An increase in the FLI and cardiometabolic risk factors in patients with CKD increases the risk of HF.

## Introduction

Metabolic dysfunction-associated steatotic liver disease (MASLD), which is replacing the previous term nonalcoholic fatty liver disease (NAFLD), has been suggested to be a risk factor not only for liver disease but also for the development and mortality of cardiovascular disease (1–3). MASLD is also a risk factor for heart failure (HF) (4, 5). A meta-analysis of 11,242,231 subjects found that MASLD was associated with a 1.5-fold higher risk of new-onset HF, regardless of the presence of hypertension, type 2 diabetes, and other cardiometabolic risk factors (4). Another study showed that patients with MASLD have a higher 10-year cumulative incidence of HF compared to those without MASLD, regardless of age and sex (5).

Chronic kidney disease (CKD) is a risk factor for the development and mortality of HF. Studies have shown that approximately 44% of patients on hemodialysis have HF (6), and about 55% of patients with HF have ≥ stage 3 CKD (7). Furthermore, a decrease in estimated glomerular filtration rate (eGFR) has been reported to be associated with an increased risk of all-cause mortality, cardiovascular mortality, and hospitalization in patients with HF (7, 8). It has also been reported that patients with both HF and CKD have a higher risk of death and hospitalization than patients with HF alone (9).

MASLD and CKD, which share common risk factors such as insulin resistance, diabetes mellitus, hypertension, and obesity, are associated with each other (10). Ultrasound-detected metabolic dysfunction-associated fatty liver disease (MAFLD) is associated with a higher risk of incident CKD (11, 12), and patients with MAFLD have been reported to have an approximately two-fold increased risk of developing end-stage renal disease (ESRD) compared with those without MAFLD (13). Furthermore, studies have shown a higher frequency of NAFLD in patients with non-diabetic CKD on hemodialysis and those with pre-dialysis CKD and an association between the severity of hepatic steatosis and decreased eGFR as well as an increased CKD stage (14).

Although both MASLD and CKD can coexist and increase the risk of HF, there have been no studies on the risk of HF according to MASLD and the type of steatotic liver disease (SLD) in patients with CKD. Therefore, in this study, we aim to investigate the risk of HF according to the co-occurrence and type of SLD in patients with CKD, using data from the National Health Insurance Service (NHIS) of South Korea.

## Methods

### Study Population

This study was conducted using data from the NHIS of South Korea. The NHIS is a national healthcare program covering the majority of South Koreans, providing information on subjects’ laboratory examination results, prescription medications, and diagnostic codes represented by the Korean Classification of Disease Seventh Revision (KCD-7) and a modified version of the International Classification of Disease Tenth Revision (ICD-10) (15, 16). The NHIS conducts regular health checkups every 2 years for individuals aged ≥ 40 years or employees. These checkups include anthropometric measurements, laboratory examinations, and self-administered health questionnaires on lifestyle behaviors such as smoking, alcohol consumption, and physical activity (17).

This study utilized data from 40% of individuals aged ≥ 20 years who underwent regular health checkups in 2012 (n=4,910,068). Among them, individuals with an eGFR of < 60 mL/min/1.73m^2^ or a previous diagnosis of ESRD (diagnostic codes V001, V003, V005) were defined as patients with CKD and included in the study (n=190,025). Individuals with a previous diagnosis of liver cancer (C22+V193, n=589), those who had undergone liver transplantation (Z944, n=181), those with a previous diagnosis of HF (n=4,824), and individuals with missing data (n=12,176) were excluded. Because outcome events were analyzed after a lag of 1 year, individuals who experienced HF within the first year were excluded (n=2,755). Individuals meeting the criteria were classified into no steatosis, MASLD, MASLD with other combined etiology, MASLD with increased alcohol intake (MetALD), alcohol-associated liver disease (ALD), specific etiology SLD, or cryptogenic SLD. The follow-up duration was from the baseline to the occurrence of HF or until December 31, 2022.

### Definitions of Disease and Measurements

Diseases were defined by diagnostic codes and laboratory examination results. The occurrence of HF was defined as having a diagnostic code I50 along with a hospitalization. Concomitant liver disease was defined as having diagnostic codes for drug-induced liver disease (K71), viral hepatitis (B15-19, B00.8, B25.1), hepatic veno-occlusive disease (I82), liver abscess (K75.0, A06.4), hemochromatosis (E83.1), Wilson’s disease (E83.0), alpha-1 antitrypsin deficiency (E88.0), autoimmune hepatitis (K75.4), primary biliary cholangitis (K74.3, K74.4), other cholangitis (K83, K83.0A), and glycogen storage disease (E74). Alcohol abuse/misuse-related disease was defined as having either diagnosis codes for alcohol abuse/misuse or other abuse- and drug-related diagnoses (E24.4, F10-F19, G31.2, G62.1, G72.1, I42.6, K29.2, K86.0, Q35.4, R78.0, T51.0, T51.8, T51.9, X65, Y15, Y57.3, Y90, Y91, Z50.2, Z71.4, Z71.2), or the diagnosis code for alcohol-related liver disease (K70). Diabetes mellitus was defined as having diagnostic codes E11-E14, along with receiving antidiabetic medications, or having a fasting glucose level of ≥ 126 mg/dL. Hypertension was defined as having diagnostic codes I10-I13, or I15, along with receiving antihypertensive medications, or having a systolic blood pressure of ≥ 140 mmHg, or a diastolic blood pressure of ≥ 90 mmHg. Dyslipidemia was defined as having diagnostic code E78, along with receiving lipid-lowering medications, or having a total cholesterol level of ≥ 240 mg/dL. Charlson Comorbidity Index (CCI) scores were calculated using the diagnosis code for each disease (18).

Blood samples were collected after an overnight fast. The Modification of Diet in Renal Disease (MDRD) equation was used to calculate eGFR (19). Urine protein levels were measured using the dipstick method, and proteinuria status was classified as negative, trace, or from 1+ to 4+.

Lifestyle behaviors, smoking status, alcohol consumption, and regular exercise were classified based on the responses obtained from self-administered health questionnaires. Smoking status was classified as never smoker, ex-smoker, and current smoker. Daily average alcohol consumption was classified as none, mild-to-moderate (< 30 g alcohol/day for males and < 20 g alcohol/day for females), heavy (30-60 g alcohol/day for males and 20-50 g alcohol/day for females), and alcoholic (≥ 60 g alcohol/day for males and ≥ 50 g alcohol/day for females). Regular exercise was defined as moderate exercise for at least 5 days per week or vigorous exercise for at least 3 days per week. Low income was defined as receiving medical aid or being in the lowest 25% income bracket.

### Surrogate Indicator and Classification of SLD

Liver ultrasonography is the gold standard for screening SLD (20); however, it is not included in the NHIS’s regular health checkups. Therefore, in this study, the fatty liver index (FLI) score, a surrogate marker for fatty liver and an accurate diagnosis of hepatic steatosis, was used to define SLD (21). The control group comprised individuals with an FLI score of < 30, and those with an FLI score of ≥ 30 were classified as having SLD.

SLD was classified according to the recommendations of the European Association for the Study of the Liver (EASL) in 2023 (1). MASLD was defined as having an FLI score ≥ 30, along with at least one cardiometabolic risk factor, ≤ mild-to moderate alcohol consumption, and the absence of concomitant liver disease and alcohol abuse/misuse-related disease. MASLD with other combined etiology was defined as having an FLI score ≥ 30, along with at least one cardiometabolic risk factor, ≤ mild-to-moderate alcohol consumption, and the presence of either concomitant liver disease or alcohol abuse/misuse-related disease. MetALD was defined as having an FLI score ≥ 30, along with at least one cardiometabolic risk factor, and heavy alcohol consumption. ALD was defined as having an FLI score ≥ 30, along with being classified as alcoholic. Specific etiology SLD was defined as having an FLI score ≥ 30, with the absence of cardiometabolic risk factors, but either consuming > mild-to-moderate alcohol consumption or having concomitant liver disease or alcohol abuse/misuse-related disease. Cryptogenic SLD was defined as having an FLI score ≥ 30, with no cardiometabolic risk factors, consuming ≤ mild-to-moderate alcohol, and the absence of both concomitant liver disease and alcohol abuse/misuse-related disease.

### Statistical Analysis

Continuous variables were presented as means ± standard deviations if they followed a normal distribution and as geometric mean (95% confidence interval) through log transformation if they did not follow a normal distribution. Categorical variables were presented as numbers (percentages). Baseline characteristics were compared using analysis of variance for continuous variables, and the chi-square test for categorical variables. The association between the type of SLD and the occurrence of HF was analyzed using Cox proportional hazard regression. Model 1 was unadjusted; model 2 was adjusted for age and sex; model 3 was adjusted for age, sex, income, smoking status, regular exercise, and CCI score; and model 4 was adjusted for age, sex, income, smoking status, regular exercise, CCI score, eGFR, and proteinuria. Stratified analyses were conducted based on sex, age, diabetes mellitus, hypertension, CCI score, and proteinuria, and Cox proportional hazard regression was conducted after adjusting for age, sex, income, smoking, regular exercise, CCI score, eGFR, and proteinuria. Additionally, individuals were classified based on their FLI, presence of cardiometabolic risk factors, and alcohol consumption level to analyze the risk of HF occurrence. *P*-values < 0.05 were considered statistically significant in all analyses. All statistical analyses were performed using SAS version 9.4 (SAS Institute, Cary, NC, USA).

### Ethical Statement

This study was approved by the Institutional Review Board of the Kangbuk Samsung Hospital (IRB no. 2024-06-043). The participants’ information was anonymized for analysis, and the consent from the study participants was waived in accordance with the Bioethics and Safety Act in South Korea.

## Results

### Baseline Characteristics

A total of 169,500 patients with CKD were analyzed based on the presence of SLD (Figure 1). Patients with MASLD and MASLD with other combined etiology were older than those without steatosis, and the proportion of male was higher in those with any type of SLD than in those without steatosis. Patients with MASLD, MASLD with other combined etiology, MetALD, and ALD had a higher BMI than those without steatosis, and the waist circumference was larger in those with all types of SLD than in those without steatosis. A higher proportion of patients with MASLD, MASLD with other combined etiology, MetALD, and ALD had hypertension, diabetes mellitus, and dyslipidemia than those without steatosis. Additionally, patients with MASLD, MASLD with other combined etiology, ALD, and specific etiology SLD had a higher proportion of those with a CCI score ≥ 2 than those without steatosis. Patients with all types of SLD had higher levels of aspartate aminotransferase, alanine aminotransferase, and gamma-glutamyl transpeptidase than patients without steatosis (Table 1).

**Figure 1.**
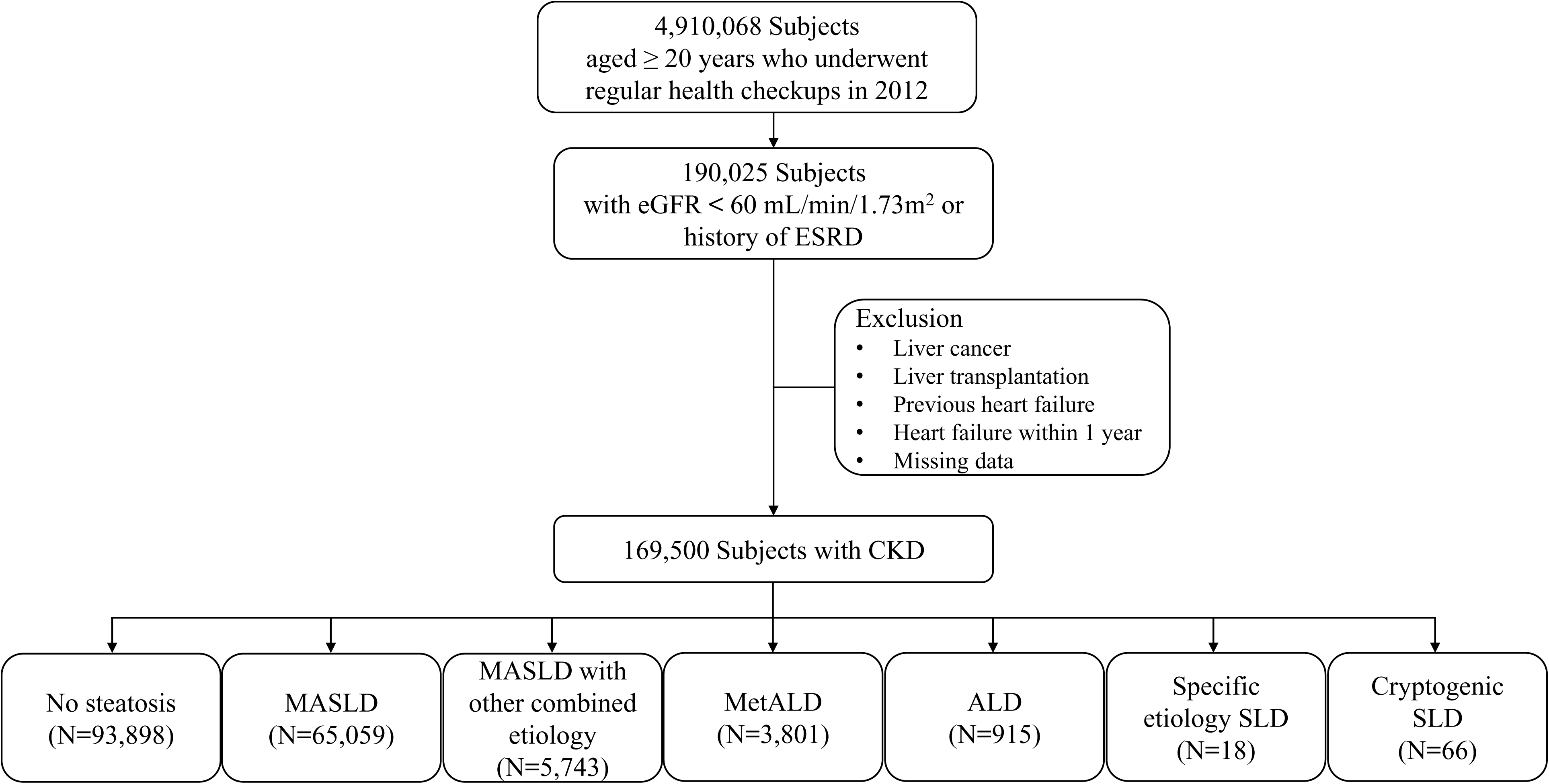
Study Subjects. ALD, alcohol-associated liver disease; CKD, chronic kidney disease; eGFR, estimated glomerular filtration rate; ESRD, end-stage renal disease; MASLD, metabolic dysfunction-associated steatotic liver disease; MetALD, MASLD with increased alcohol intake; SLD, steatotic liver disease.

**Table 1.**
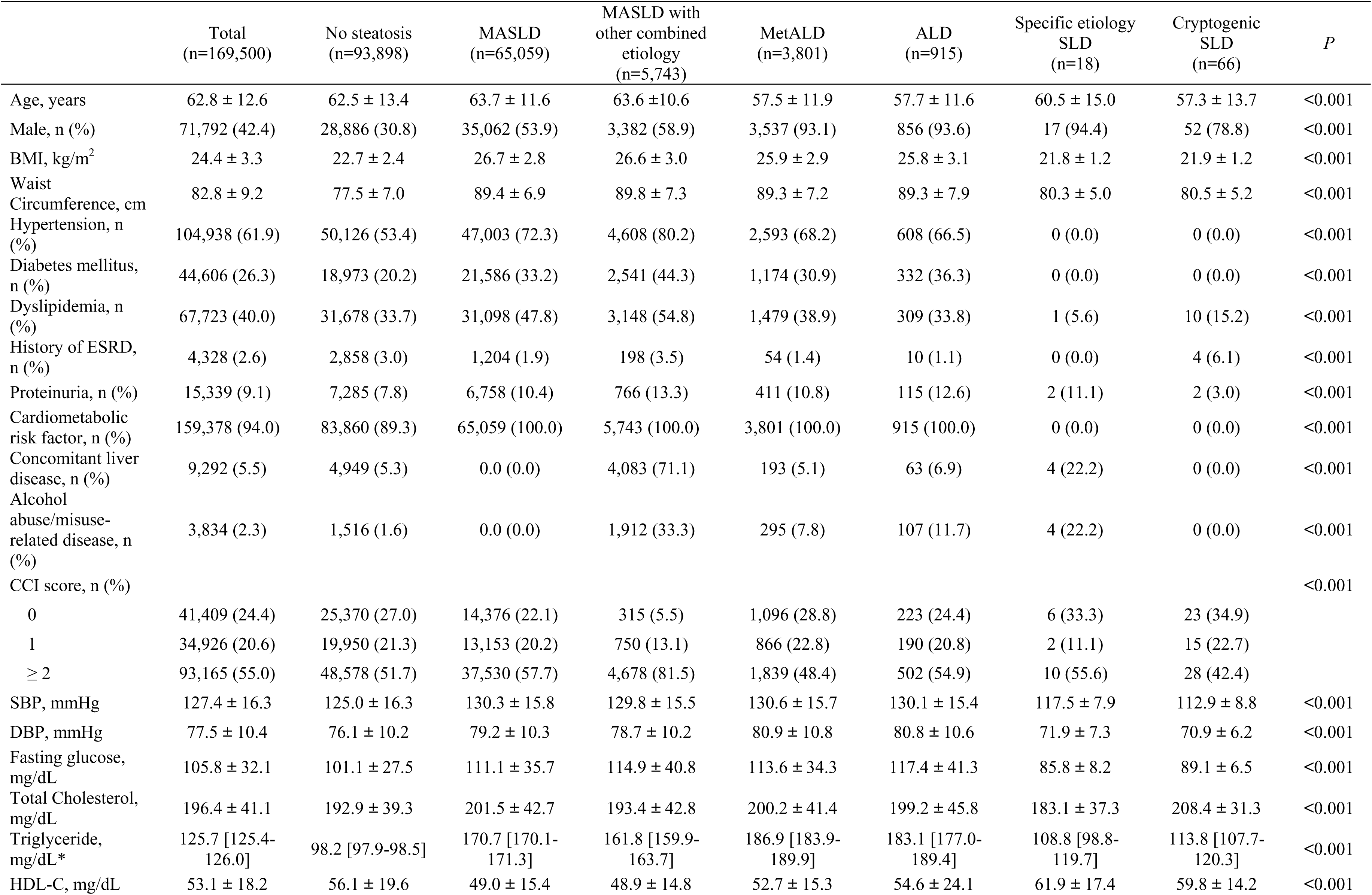
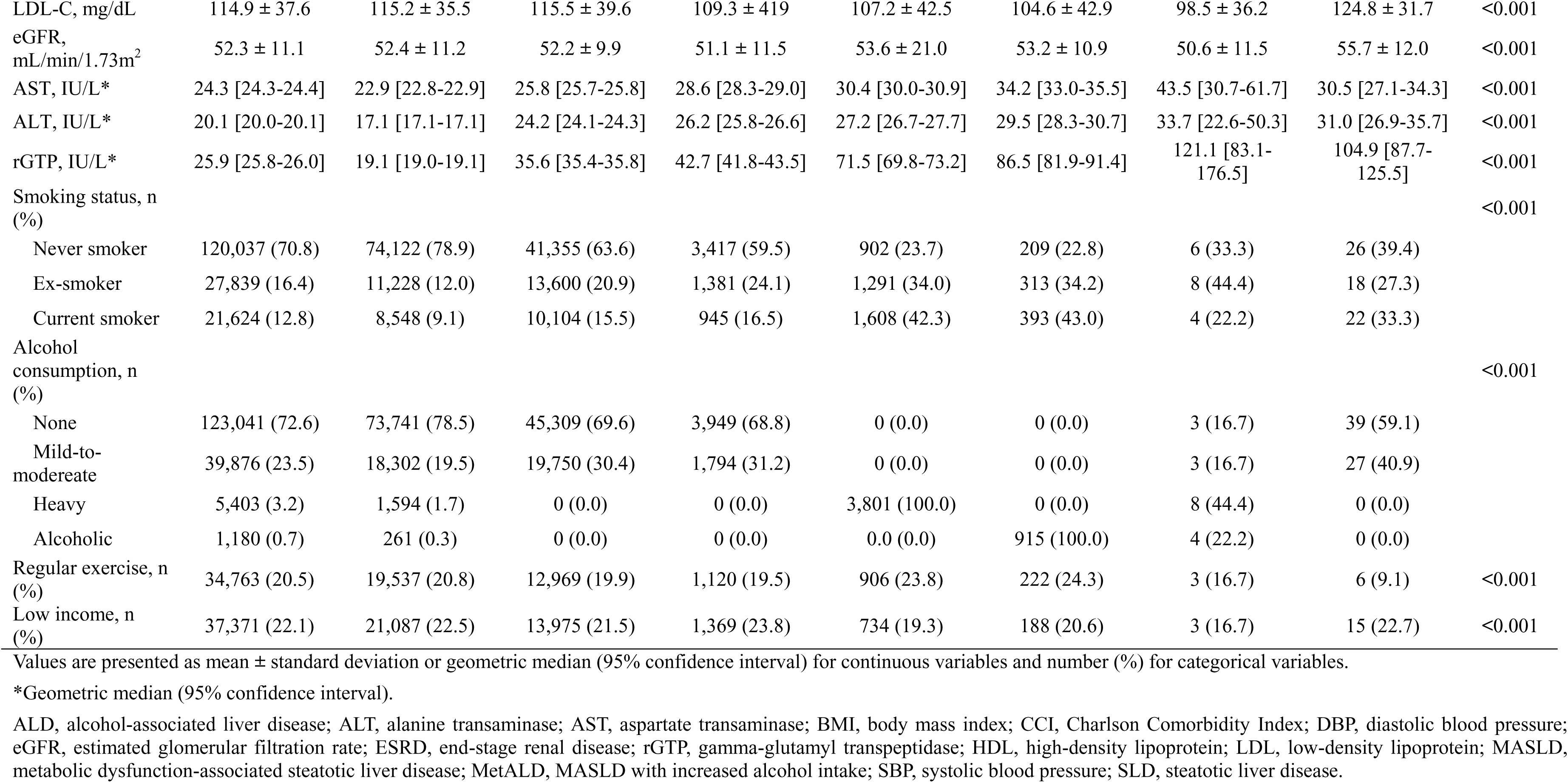
Baseline Characteristics of the Study Population.

### Risk of HF According to SLD

During the median follow-up period of 9.2 years, a total of 29,555 (17.4%) HF cases occurred. HF occurred in 12,698 (19.5%) patients with MASLD, 1,390 (24.2%) with MASLD with other combined etiology, 5 (27.8%) with specific etiology SLD, and 15 (22.7%) with cryptogenic SLD, showing a higher incidence of HF in these patients than in those without steatosis (14,752 [15.7%]).

When analyzed using the Cox proportional hazards model adjusted for age, sex, income, smoking status, regular exercise, CCI score, eGFR, and proteinuria, MASLD (hazard ratio [95% confidence interval]: 1.133 [1.106, 1.161]), MASLD with other combined etiology (1.276 [1.207, 1.350]), and cryptogenic SLD (1.817 [1.095, 3.015]) were associated with an increased risk of HF compared with no steatosis (Table 2).

**Table 2.** Risk of Heart Failure according to Type of Steatotic Liver Disease.

|  | Event, n (%) | Incidence rate,<br>1,000 p-y | Model 1 | Model 2 | Model 3 | Model 4 |
| --- | --- | --- | --- | --- | --- | --- |
|  |  |  | HR [95% CI] |  |  |  |
| No steatosis | 14,752 (15.7) | 19.0 | 1 (Ref.) | 1 (Ref.) | 1 (Ref.) | 1 (Ref.) |
| MASLD | 12,698 (19.5) | 23.9 | 1.270 [1.240, 1.300] | 1.176 [1.148, 1.205] | 1.133 [1.106, 1.161] | 1.133 [1.106, 1.161] |
| MASLD with other<br>combined etiology | 1,390 (24.2) | 31.1 | 1.669 [1.580, 1.764] | 1.581 [1.496, 1.670] | 1.264 [1.196, 1.337] | 1.276 [1.207, 1.350] |
| MetALD | 553 (14.6) | 17.3 | 0.912 [0.837, 0.992] | 1.079 [0.990, 1.177] | 1.023 [0.939, 1.116] | 1.057 [0.969, 1.152] |
| ALD | 142 (15.5) | 18.8 | 0.990 [0.839, 1.167] | 1.184 [1.003, 1.398] | 1.101 [0.933, 1.300] | 1.120 [0.948, 1.322] |
| Specific etiology SLD | 5 (27.8) | 33.8 | 1.793 [0.746, 4.309] | 1.706 [0.710, 4.100] | 1.836 [0.764, 4.412] | 2.003 [0.834, 4.815] |
| Cryptogenic SLD | 15 (22.7) | 27.7 | 1.469 [0.886, 2.438] | 1.754 [1.057, 2.910] | 1.625 [0.979, 2.697] | 1.817 [1.095, 3.015] |

Model 1: unadjusted. Model 2: adjusted for age and sex. Model 3: adjusted for age, sex, income, smoking status, regular exercise, and CCI score. Model 4: adjusted for age, sex, income, smoking status, regular exercise, CCI score, eGFR, and proteinuria.
ALD, alcohol-associated liver disease; CI, confidence interval; HF, heart failure; HR, hazard ratio; MASLD, metabolic dysfunction-associated steatotic liver disease; MetALD, MASLD with increased alcohol intake; p-y, person-year; SLD, steatotic liver disease.

### Stratified Analyses of Risk of HF According to SLD

A stratified subgroup analysis was conducted according to sex (male vs. female), age (< 65 vs. ≥ 65 years), presence of diabetes mellitus, presence of hypertension, CCI score (0 vs. 1 vs. ≥ 2), and presence of proteinuria (Figure 2). The association between MASLD, MASLD with other combined etiology, and the occurrence of HF, compared with no steatosis, was more prominent in female, younger age, those without hypertension, and those without proteinuria. The association between cryptogenic SLD and the occurrence of HF, compared with no steatosis, was more prominent in younger age, those without hypertension, and those with proteinuria. MetALD was also associated with the occurrence of HF in female. In addition, specific etiology SLD was associated with the occurrence of HF in those without hypertension and those with proteinuria.

**Figure 2.**
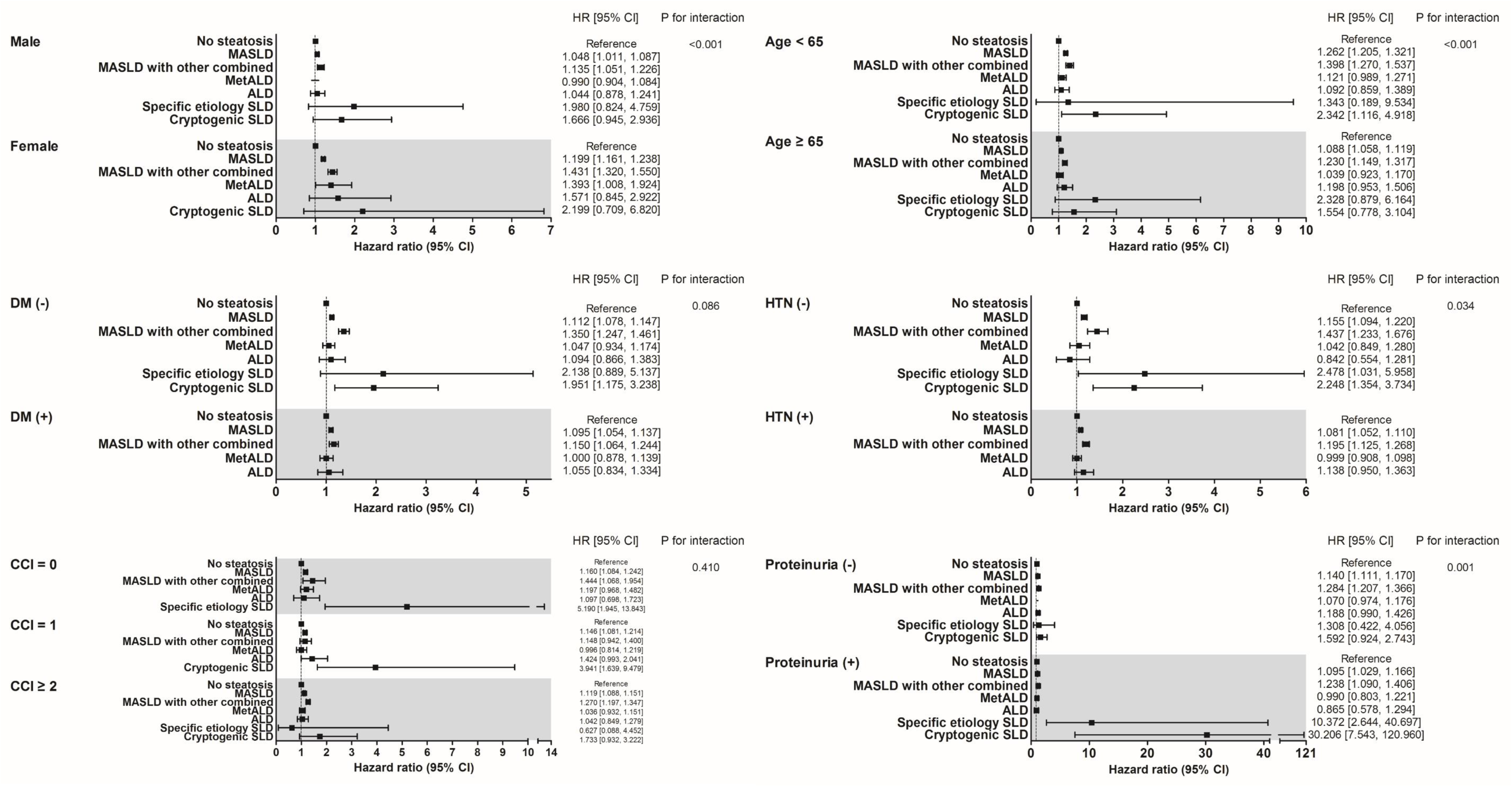
Stratified Analyses of Risk of HF According to SLD. Adjusted for age, sex, income, smoking statis, regular exercise, CCI, eGFR, and proteinuria. ALD, alcohol-associated liver disease; CCI, Charlson Comorbidity Index; CI, confidence interval; DM, diabetes mellitus; eGFR, estimated glomerular filtration rate; HF, heart failure; HTN, hypertension; MASLD, metabolic dysfunction-associated steatotic liver disease; MetALD, MASLD with increased alcohol intake; SLD, steatotic liver disease.

### Risk of HF According to FLI, Cardiometabolic Risk Factors and Alcohol Consumption

We analyzed the risk of HF according to FLI (< 30 and ≥ 30), presence or absence of cardiometabolic risk factors, and levels of alcohol consumption (none, mild-to-moderate, heavy, and alcoholic) (Table 3). The groups with an FLI < 30 and cardiometabolic risk factors, FLI ≥ 30 without cardiometabolic risk factors, and FLI ≥ 30 with cardiometabolic risk factors all had a higher incidence of HF, regardless of the level of alcohol consumption, than the group with FLI < 30, no cardiometabolic risk factors, and none alcohol consumption. When analyzed using the Cox proportional hazards model adjusted for age, sex, income, smoking status, regular exercise, CCI score, eGFR, and proteinuria, the groups with an FLI < 30 and cardiometabolic risk factors, FLI ≥ 30 with no cardiometabolic risk factors and none alcohol consumption, and FLI ≥ 30 with cardiometabolic risk factors all were associated with a higher incidence of HF than the group with an FLI < 30, no cardiometabolic risk factors, and none alcohol consumption. Similar results were observed when patients with concomitant liver disease and alcohol abuse/misuse-related disease were excluded (Supplementary Table 1).

**Table 3.** Risk of Heart Failure according to Fatty Liver Index, Cardiometabolic Risk Factors and Alcohol Consumption.

| FLI | Cardiometabolic risk factor | Alcohol consumption | Event, n (%) | Duration, p-y | Incidence rate, 1000 p-y | Model 1 | Model 2 | Model 3 | Model 4 |
| --- | --- | --- | --- | --- | --- | --- | --- | --- | --- |
|  |  |  |  |  |  | HR [95% CI] |  |  |  |
| < 30 | No | None | 426 (6.2) | 61041.4 | 7.0 | 1.000 (Reference) | 1.000 (Reference) | 1.000 (Reference) | 1.000 (Reference) |
|  |  | Mild-to-moderate | 113 (3.8) | 26969.2 | 4.2 | 0.598 [0.486, 0.736] | 0.787 [0.640, 0.969] | 0.785 [0.637, 0.966] | 0.813 [0.660, 1.000] |
|  |  | Heavy | 8 (3.6) | 1998.4 | 4.0 | 0.573 [0.285, 1.153] | 0.728 [0.362, 1.464] | 0.754 [0.375, 1.517] | 0.752 [0.373, 1.513] |
|  |  | Alcoholic | 2 (4.8) | 365.0 | 5.5 | 0.792 [0.198, 3.178] | 0.656 [0.163, 2.630] | 0.599 [0.149, 2.405] | 0.633 [0.158, 2.540] |
|  | Yes | None | 12101 (18.1) | 543756.8 | 22.3 | 3.258 [2.958, 3.588] | 1.643 [1.491, 1.811] | 1.438 [1.305, 1.585] | 1.356 [1.230, 1.495] |
|  |  | Mild-to-moderate | 1,886 (12.3) | 131240.3 | 14.4 | 2.083 [1.875, 2.314] | 1.361 [1.224, 1.513] | 1.228 [1.105, 1.366] | 1.186 [1.067, 1.319] |
|  |  | Heavy | 184 (13.4) | 11345.8 | 16.2 | 2.368 [1.992, 2.815] | 1.523 [1.280, 1.811] | 1.316 [1.106, 1.566] | 1.259 [1.058, 1.498] |
|  |  | Alcoholic | 32 (14.6) | 1685.6 | 19.0 | 2.805 [1.959, 4.018] | 1.841 [1.285, 2.637] | 1.617 [1.129, 2.317] | 1.677 [1.170, 2.403] |
| ≥ 30 | No | None | 13 (31.0) | 330.6 | 39.3 | 5.802 [3.341, 10.074] | 3.237 [1.864, 5.622] | 2.740 [1.577, 4.758] | 2.842 [1.636, 4.936] |
|  |  | Mild-to-moderate | 3 (10.0) | 254.4 | 11.8 | 1.724 [0.554, 5.368] | 1.373 [0.441, 4.273] | 1.065 [0.342, 3.317] | 1.213 [0.390, 3.777] |
|  |  | Heavy | 3 (37.5) | 73.1 | 41.1 | 5.881 [1.889, 18.306] | 2.296 [0.737, 7.149] | 2.489 [0.799, 7.751] | 2.591 [0.832, 8.066] |
|  |  | Alcoholic | 1 (25.0) | 32.1 | 31.2 | 4.636 [0.652, 32.988] | 4.826 [0.678, 34.342] | 4.347 [0.611, 30.940] | 3.681 [0.517, 26.203] |
|  | Yes | None | 10,977 (22.3) | 393179.2 | 27.9 | 4.114 [3.735, 4.533] | 1.975 [1.792, 2.177] | 1.645 [1.492, 1.814] | 1.554 [1.410, 1.714] |
|  |  | Mild-to-moderate | 3,111 (14.4) | 182347.5 | 17.1 | 2.483 [2.244, 2.748] | 1.519 [1.371, 1.683] | 1.325 [1.196, 1.468] | 1.295 [1.168, 1.435] |
|  |  | Heavy | 553 (14.5) | 31994.2 | 17.3 | 2.517 [2.219, 2.856] | 1.589 [1.399, 1.805] | 1.342 [1.181, 1.525] | 1.327 [1.168, 1.508] |
|  |  | Alcoholic | 142 (15.5) | 7570.3 | 18.8 | 2.734 [2.261, 3.306] | 1.742 [1.439, 2.107] | 1.443 [1.192, 1.746] | 1.404 [1.160, 1.700] |

Model 1: unadjusted. Model 2: adjusted for age and sex. Model 3: adjusted for age, sex, income, smoking status, regular exercise, and CCI score. Model 4: adjusted for age, sex, income, smoking status, regular exercise, CCI score, eGFR, and proteinuria.
CI, confidence interval; FLI, fatty liver index; HF, heart failure; HR, hazard ratio.

## Discussion

When patients with CKD were classified according to SLD, the incidence of HF was higher in patients with MASLD, MASLD with other combined etiology, and cryptogenic SLD than in those without steatosis. The association between MASLD, MASLD with other combined etiology, and HF was more prominent in women, younger age, those without hypertension, and those without proteinuria, and the association between cryptogenic SLD and HF was more prominent in younger age, those without hypertension, and those with proteinuria. In patients with CKD, an increase in the FLI and the presence of cardiometabolic factors were associated with an increased risk of HF.

The EASL classifies SLD into the following categories: MASLD, MetALD, ALD, specific etiology SLD, and cryptogenic SLD. A notable change in the definition of SLD is the replacement of the term NAFLD with MASLD, which is defined as hepatic steatosis with at least one of five cardiometabolic risk factors. This change was made to provide a clearer understanding of the etiology of the disease and avoid the stigmatization of patients (1). MASLD, similar to NAFLD, is associated with a higher risk of cardiovascular disease (22, 23). Another study comparing patients with NAFLD and MASLD found that patients with MASLD are older and had a slightly higher mortality risk, although no significant differences in mortality were observed between patients with NAFLD and those with MASLD (24).

SLD contributes to the development of HF through multiple mechanisms (25–31). Early stage functional alterations such as endothelial dysfunction and hyperreactivity to vasoconstrictors (25–27), along with early stage fibrosis and hepatocellular enlargement caused by lipid accumulation, can lead to portal hypertension, thus affecting the preload reserve (28). Additionally, advanced glycation end-products (AGEs) resulting from insulin resistance can activate fibrosis-inducing factors (29) and contribute to the formation of reactive oxygen species, which can influence myocardial cell fibrosis (30). Furthermore, the upregulated sympathetic nervous system in MASLD can influence the renin-angiotensin-aldosterone system (RAAS), contributing to cardiac fibrosis (31). Mechanisms leading to HF in patients with SLD can also manifest in those with CKD. CKD induces hemodynamic changes and causes salt and water retention, leading to venous congestion, myocardial cell lengthening, and eccentric left ventricular (LV) remodeling. In addition, CKD promotes myocardial cell thickening and concentric LV remodeling by increasing systemic arterial resistance and enhancing large-vessel compliance through processes such as vascular calcification. These preload- and afterload-related factors of CKD lead to myocardial hypertrophy and myocyte ischemia, causing interstitial myocardial cell fibrosis, ultimately resulting in HF (32). In addition to these hemodynamic factors, oxidative stress and inflammation in CKD increase myocardial fibrosis, thereby affecting the occurrence of HF (33, 34). Furthermore, in patients with CKD, the RAAS is excessively activated, resulting in angiotensin II and aldosterone inducing myocardial cell hypertrophy and fibrosis independent of afterload (35, 36). The activated sympathetic nervous system in CKD (37) increases renin secretion (38) and also contributes to interstitial myocardial cell fibrosis (39). Given that SLD and CKD contribute to the occurrence of HF through similar and distinct mechanisms, the presence of SLD in patients with CKD may increase the risk of HF compared with those without SLD. Indeed, in this study, patients with CKD and MASLD, MASLD with other combined etiology, and cryptogenic SLD were found to be associated with a higher incidence of HF than those without steatosis, and patients with CKD with a higher FLI were associated with a higher incidence of HF than those with a lower FLI. In particular, the association between MASLD, MASLD with other combined etiology, and HF was more prominent in women than in men, which is thought to be due to activation of inflammatory mechanisms due to a loss of estrogen in old age (40). Additionally, the association between MASLD, MASLD with other combined etiology, and HF was more prominent in subjects without proteinuria, which is associated with diabetes mellitus, obesity, and insulin resistance and is one of the risk factors for heart failure. In this study, urine protein levels were measured using the dipstick method, which has low sensitivity (41). Additionally, medications administered for proteinuria may have affected the results.

In this study, MASLD and the presence of cardiometabolic risk factors were associated with a higher incidence of HF. However, MetALD and ALD did not increase the incidence of HF, nor did increased alcohol consumption. This suggests that in patients with CKD and SLD, not only SLD itself but also cardiometabolic risk factors such as obesity, hyperglycemia, hypertension, and dyslipidemia influence the occurrence of HF. Furthermore, while alcohol is known to have a direct toxic effect on the myocardium through high oxidative stress and apoptosis and to increase norepinephrine levels, adversely affecting the cardiovascular system (42, 43), conflicting results regarding mild-to-moderate alcohol intake and LV function (44, 45) suggest that alcohol might not have had a significant impact on the occurrence of HF in patients with both CKD and SLD. In addition, alcohol consumption was based on self-administered questionnaires, which may have led to inaccuracies. This study did not account for the type or duration of alcohol consumption, which may have influenced the results. The smaller number of subjects with MetALD and ALD compared to MASLD may have influenced the results. In addition, not only MASLD but also MASLD with other combined etiology, in which MASLD was accompanied by concomitant liver disease or alcohol abuse/misuse-related disease, was associated with a high incidence of HF. Previous studies have reported that HF is commonly found in hemochromatosis and that cardiomyopathy occurs in patients with liver abscess (46, 47). In addition, patients with alpha-1 antitrypsin deficiency had a high risk of hospitalization for HF and HF- specific mortality due to remodeling of cardiac tissue resulting from the alpha-1 antitrypsin deficiency (48). Considering the results of these previous studies and the present study, hepatic steatosis and cardiometabolic risk factors, as well as concomitant liver diseases such as hemochromatosis and liver abscess, are thought to be factors that may contribute to the development of HF.

This study had several limitations. This study utilized data from individuals who underwent national health examinations (66.0% in 2009 (49)), potentially leading to selection bias. Furthermore, this study used the FLI to screen for hepatic steatosis, which has limitations in accurately quantifying hepatic steatosis and distinguishing between simple steatosis and steatohepatitis. Additionally, the occurrence of HF was defined solely based on diagnostic codes without echocardiography results. This study also has a limitation in that it did not take into account medications for hypertension, diabetes mellitus, and hyperlipidemia, which may affect the development of HF. Furthermore, CKD was defined based on eGFR calculated using creatinine values, which may lead to overestimation in patients with low muscle mass (50). However, to our knowledge, this study is the first to analyze the risk of HF associated with SLD in patients with CKD, thus demonstrating its clinical significance. Additionally, a strength of this study is that it includes not only MASLD and MetALD but also specific etiology SLD, cryptogenic SLD based on the classification criteria recently presented by EASL. Recently, several medications have been reported to reduce mortality due to HF; however, their use is limited in patients with CKD. Taking this into consideration, the findings of the current study suggesting an increased occurrence of HF in patients with CKD and SLD may contribute to reducing the incidence of HF in patients with CKD.

In this large population-based cohort study involving patients with CKD, the presence of MASLD, MASLD with other combined etiology, and cryptogenic SLD was associated with a higher incidence of HF than the absence of steatosis. Moreover, an increase in the FLI and the presence of cardiometabolic risk factors increased the risk of HF. Considering the high mortality rate associated with HF, managing SLD and cardiometabolic risk factors in patients with CKD is crucial.

## Data Availability

Some or all datasets generated during and/or analyzed during the current study are not publicly available but are available from the corresponding author on reasonable request.

## Acknowledgments

We would like to thank Gyuna Lee for her assistance with statistical analysis.

## References

1. Rinella ME, Lazarus JV, Ratziu V, Francque SM, Sanyal AJ, Kanwal F, et al. A multisociety Delphi consensus statement on new fatty liver disease nomenclature. Hepatology. 2023;78(6):1966–86.

2. Loomba R, Friedman SL, Shulman GI. Mechanisms and disease consequences of nonalcoholic fatty liver disease. Cell. 2021;184(10):2537–64.

3. Targher G, Byrne CD, Lonardo A, Zoppini G, Barbui C. Non-alcoholic fatty liver disease and risk of incident cardiovascular disease: A meta-analysis. J Hepatol. 2016;65(3):589–600.

4. Mantovani A, Petracca G, Csermely A, Beatrice G, Bonapace S, Rossi A, et al. Non-alcoholic fatty liver disease and risk of new-onset heart failure: an updated meta-analysis of about 11 million individuals. Gut. 2022 Jul;gutjnl-2022-327672.

5. Roderburg C, Krieg S, Krieg A, Vaghiri S, Mohr R, Konrad M, et al. Non-Alcoholic Fatty Liver Disease (NAFLD) and risk of new-onset heart failure: a retrospective analysis of 173,966 patients. Clin Res Cardiol. 2023;112(10):1446–53.

6. House AA, Wanner C, Sarnak MJ, Pina IL, McIntyre CW, Komenda P, et al. Heart failure in chronic kidney disease: conclusions from a Kidney Disease: Improving Global Outcomes (KDIGO) Controversies Conference. Kidney Int. 2019;95(6):1304–17.

7. McAlister FA, Ezekowitz J, Tarantini L, Squire I, Komajda M, Bayes-Genis A, et al. Renal dysfunction in patients with heart failure with preserved versus reduced ejection fraction: impact of the new Chronic Kidney Disease-Epidemiology Collaboration Group formula. Circ Heart Fail. 2012;5(3):309–14.

8. Hillege HL, Nitsch D, Pfeffer MA, Swedberg K, McMurray JJ, Yusuf S, et al. Renal function as a predictor of outcome in a broad spectrum of patients with heart failure. Circulation. 2006;113(5):671–8.

9. Smith DH, Thorp ML, Gurwitz JH, McManus DD, Goldberg RJ, Allen LA, et al. Chronic kidney disease and outcomes in heart failure with preserved versus reduced ejection fraction: the Cardiovascular Research Network PRESERVE Study. Circ Cardiovasc Qual Outcomes. 2013;6(3):333–42.

10. Cheung A, Ahmed A. Nonalcoholic Fatty Liver Disease and Chronic Kidney Disease: A Review of Links and Risks. Clin Exp Gastroenterol. 2021;14:457–65.

11. Liang Y, Chen H, Liu Y, Hou X, Wei L, Bao Y, et al. Association of MAFLD With Diabetes, Chronic Kidney Disease, and Cardiovascular Disease: A 4.6-Year Cohort Study in China. J Clin Endocrinol Metab. 2022;107(1):88-97.

12. Wei S, Song J, Xie Y, Huang J, Yang J. Metabolic dysfunction-associated fatty liver disease can significantly increase the risk of chronic kidney disease in adults with type 2 diabetes. Diabetes Res Clin Pract. 2023;197:110563.

13. Chen S, Pang J, Huang R, Xue H, Chen X. Association of MAFLD with end-stage kidney disease: a prospective study of 337,783 UK Biobank participants. Hepatol Int. 2023;17(3):595–605.

14. Behairy MA, Sherief AF, Hussein HA. Prevalence of non-alcoholic fatty liver disease among patients with non-diabetic chronic kidney disease detected by transient elastography. Int Urol Nephrol. 2021;53(12):2593–601.

15. Lee J, Lee JS, Park SH, Shin SA, Kim K. Cohort Profile: The National Health Insurance Service-National Sample Cohort (NHIS-NSC), South Korea. Int J Epidemiol. 2017;46(2):e15.

16. Cho SW, Kim JH, Choi HS, Ahn HY, Kim MK, Rhee EJ. Big Data Research in the Field of Endocrine Diseases Using the Korean National Health Information Database. Endocrinol Metab (Seoul). 2023;38(1):10–24.

17. Lee YH, Han K, Ko SH, Ko KS, Lee KU, Taskforce Team of Diabetes Fact Sheet of the Korean Diabetes A. Data Analytic Process of a Nationwide Population-Based Study Using National Health Information Database Established by National Health Insurance Service. Diabetes Metab J. 2016;40(1):79–82.

18. Glasheen WP, Cordier T, Gumpina R, Haugh G, Davis J, Renda A. Charlson Comorbidity Index: ICD-9 Update and ICD-10 Translation. Am Health Drug Benefits. 2019;12(4):188–97.

19. Levey AS, Bosch JP, Lewis JB, Greene T, Rogers N, Roth D. A more accurate method to estimate glomerular filtration rate from serum creatinine: a new prediction equation. Modification of Diet in Renal Disease Study Group. Ann Intern Med. 1999;130(6):461–70.

20. Chalasani N, Younossi Z, Lavine JE, Diehl AM, Brunt EM, Cusi K, et al. The diagnosis and management of non-alcoholic fatty liver disease: practice guideline by the American Gastroenterological Association, American Association for the Study of Liver Diseases, and American College of Gastroenterology. Gastroenterology. 2012;142(7):1592–609.

21. Bedogni G, Bellentani S, Miglioli L, Masutti F, Passalacqua M, Castiglione A, et al. The Fatty Liver Index: a simple and accurate predictor of hepatic steatosis in the general population. BMC Gastroenterol. 2006;6:33.

22. Lee HH, Lee HA, Kim EJ, Kim HY, Kim HC, Ahn SH, et al. Metabolic dysfunction-associated steatotic liver disease and risk of cardiovascular disease. Gut. 2024;73(3):533–40.

23. Moon JH, Jeong S, Jang H, Koo BK, Kim W. Metabolic dysfunction-associated steatotic liver disease increases the risk of incident cardiovascular disease: a nationwide cohort study. EClinicalMedicine. 2023;65:102292.

24. Younossi ZM, Paik JM, Stepanova M, Ong J, Alqahtani S, Henry L. Clinical profiles and mortality rates are similar for metabolic dysfunction-associated steatotic liver disease and non-alcoholic fatty liver disease. J Hepatol. 2024;80(5):694–701.

25. Pasarin M, La Mura V, Gracia-Sancho J, Garcia-Caldero H, Rodriguez-Vilarrupla A, Garcia-Pagan JC, et al. Sinusoidal endothelial dysfunction precedes inflammation and fibrosis in a model of NAFLD. PLoS One. 2012;7(4):e32785.

26. van der Graaff D, Chotkoe S, De Winter B, De Man J, Casteleyn C, Timmermans JP, et al. Vasoconstrictor antagonism improves functional and structural vascular alterations and liver damage in rats with early NAFLD. JHEP Rep. 2022;4(2):100412.

27. Van der Graaff D, Kwanten WJ, Couturier FJ, Govaerts JS, Verlinden W, Brosius I, et al. Severe steatosis induces portal hypertension by systemic arterial hyporeactivity and hepatic vasoconstrictor hyperreactivity in rats. Lab Invest. 2018;98(10):1263–75.

28. Baffy G. Origins of Portal Hypertension in Nonalcoholic Fatty Liver Disease. Dig Dis Sci. 2018;63(3):563–76.

29. Daoud S, Schinzel R, Neumann A, Loske C, Fraccarollo D, Diez C, et al. Advanced glycation endproducts: activators of cardiac remodeling in primary fibroblasts from adult rat hearts. Mol Med. 2001;7(8):543–51.

30. Yan SD, Schmidt AM, Anderson GM, Zhang J, Brett J, Zou YS, et al. Enhanced cellular oxidant stress by the interaction of advanced glycation end products with their receptors/binding proteins. J Biol Chem. 1994;269(13):9889–97.

31. Sigala B, McKee C, Soeda J, Pazienza V, Morgan M, Lin CI, et al. Sympathetic nervous system catecholamines and neuropeptide Y neurotransmitters are upregulated in human NAFLD and modulate the fibrogenic function of hepatic stellate cells. PLoS One. 2013;8(9):e72928.

32. Glassock RJ, Pecoits-Filho R, Barberato SH. Left ventricular mass in chronic kidney disease and ESRD. Clin J Am Soc Nephrol. 2009;4 Suppl 1:S79–91.

33. Paoletti E, Bellino D, Cassottana P, Rolla D, Cannella G. Left ventricular hypertrophy in nondiabetic predialysis CKD. Am J Kidney Dis. 2005;46(2):320–7.

34. Gupta J, Dominic EA, Fink JC, Ojo AO, Barrows IR, Reilly MP, et al. Association between Inflammation and Cardiac Geometry in Chronic Kidney Disease: Findings from the CRIC Study. PLoS One. 2015;10(4):e0124772.

35. Schunkert H, Sadoshima J, Cornelius T, Kagaya Y, Weinberg EO, Izumo S, et al. Angiotensin II-induced growth responses in isolated adult rat hearts. Evidence for load-independent induction of cardiac protein synthesis by angiotensin II. Circ Res. 1995;76(3):489–97.

36. Steigerwalt S, Zafar A, Mesiha N, Gardin J, Provenzano R. Role of aldosterone in left ventricular hypertrophy among African-American patients with end-stage renal disease on hemodialysis. Am J Nephrol. 2007;27(2):159–63.

37. Converse RL, Jr., Jacobsen TN, Toto RD, Jost CM, Cosentino F, Fouad-Tarazi F, et al. Sympathetic overactivity in patients with chronic renal failure. N Engl J Med. 1992;327(27):1912–8.

38. DiBona GF. Physiology in perspective: The Wisdom of the Body. Neural control of the kidney. Am J Physiol Regul Integr Comp Physiol. 2005;289(3):R633–41.

39. Perlini S, Palladini G, Ferrero I, Tozzi R, Fallarini S, Facoetti A, et al. Sympathectomy or doxazosin, but not propranolol, blunt myocardial interstitial fibrosis in pressure-overload hypertrophy. Hypertension. 2005;46(5):1213–8.

40. Regitz-Zagrosek V. Sex and Gender Differences in Heart Failure. Int J Heart Fail. 2020;2(3):157–81.

41. Panta P, Techakehakij W. Diagnostic accuracy of a urine dipstick for detecting albuminuria in hypertensive patients. F1000Res. 2020;9:1244.

42. Chen DB, Wang L, Wang PH. Insulin-like growth factor I retards apoptotic signaling induced by ethanol in cardiomyocytes. Life Sci. 2000;67(14):1683–93.

43. Adams MA, Hirst M. Metoprolol suppresses the development of ethanol-induced cardiac hypertrophy in the rat. Can J Physiol Pharmacol. 1990;68(5):562–7.

44. Gemes K, Janszky I, Strand LB, Laszlo KD, Ahnve S, Vatten LJ, et al. Light-moderate alcohol consumption and left ventricular function among healthy, middle-aged adults: the HUNT study. BMJ Open. 2018;8(5):e020777.

45. Park SK, Moon K, Ryoo JH, Oh CM, Choi JM, Kang JG, et al. The association between alcohol consumption and left ventricular diastolic function and geometry change in general Korean population. Eur Heart J Cardiovasc Imaging. 2018;19(3):271–8.

46. Seferovic PM, Polovina M, Bauersachs J, Arad M, Ben Gal T, Lund LH, et al. Heart failure in cardiomyopathies: a position paper from the Heart Failure Association of the European Society of Cardiology. Eur J Heart Fail. 2019;21(5):553–76.

47. Erem AS, Krapivina A, Braverman TS, Allamaneni SS. Serratia Liver Abscess Infection and Cardiomyopathy in a Patient with Diabetes Mellitus: A Case Report and Review of the Literature. Am J Case Rep. 2019;20:1343–9.

48. Winther SV, Landt EM, Nordestgaard BG, Seersholm N, Dahl M. alpha(1)- Antitrypsin deficiency associated with increased risk of heart failure. ERJ Open Res. 2023;9(5).

49. Moon SJ, Rhee EJ, Jung JH, Han KD, Kim SR, Lee WY, et al. Independent Impact of Diabetes on the Severity of Coronavirus Disease 2019 in 5,307 Patients in South Korea: A Nationwide Cohort Study. Diabetes Metab J. 2020;44(5):737–46.

50. Groothof D, Post A, Polinder-Bos HA, Erler NS, Flores-Guerrero JL, Kootstra-Ros JE, et al. Muscle mass and estimates of renal function: a longitudinal cohort study. J Cachexia Sarcopenia Muscle. 2022;13(4):2031–43.

